# Quantifying Health Gains and Health System Expenditure Impacts of Eliminating Indoor Mould in Homes

**DOI:** 10.1101/2025.04.13.25325769

**Authors:** Yuxi Li, Shiva Raj Mishra, Tim Wilson, Rebecca Bentley, Tony Blakely

**Author notes:** Corresponding author: Yuxi Li Centre for Health Policy, Melbourne School of Population and Global Health, University of Melbourne, 207 Bouverie Street, Melbourne VIC 3000, Australia. YL † led the literature searches, final specification of model, and final drafting of paper. SRM † led initial literature searches, initial specification of model, and initial drafting of paper. TW led the build and running of the simulation models, and contributed to final drafting of paper. RB ‡ co-led overall conceptualisation of paper, led the securing of funding, had oversight of all inputs of housing- and mould-related variables, and contributed to final drafting as co-senior author. TB ‡ co-led overall conceptualisation of paper, contributed to securing funding, had oversight of all inputs to modelling, and contributed to final drafting as co-senior author. Joint first author. Joint senior author.

## Abstract

**Objective:** To quantify the prospective health benefits, cost savings, and income gains of eradicating indoor mould in Australia.

**Methods:** We used a proportional multistate lifetable model to estimate the impact of removing moderate (16.5%) and severe (11.0%) indoor mould exposure, incorporating its causal links to asthma incidence, chronic respiratory disease severity, and lower respiratory tract infections. Outcomes included health-adjusted life years (HALYs), healthcare costs, and income effects, discounted at 3% annually.

**Results:** Mould eradication yielded 4,190 HALYs gained (95% UI: 1,920 to 7,350) per million people alive in 2021 over 20 years (or about 1.5 healthy days per person). The most socioeconomically deprived quintile experienced 1.66 times the benefit of the least deprived. Health expenditure decreased by US$78.0 million per million people (95% UI: 34.6 to 138), while income increased by US$116 million (95% UI: 48.7 to 219), representing 0.5%–2.1% of annual health spending and 0.08%–0.36% of GDP.

**Conclusions:** Mould eradication offers health gains comparable to other leading public health interventions and triple those from eliminating cold housing. Further research should reduce uncertainty in risk estimates and evaluate cost-effective housing solutions.

## 1. INTRODUCTION

Indoor dampness and mould are pervasive issues, impacting an estimated 5% to 30% of homes in cold climates and an 10% to 60% in regions with moderate to warm climates ^1^. While advancements in housing construction and public awareness may reduce future risk, the intensifying influence of climate change, particularly through increased heavy rainfall, flooding, and shifting humidity patterns, are expected to amplify mould- related problems in the existing housing stock. These environmental pressures highlight an urgent need for adaptation and mitigation strategies.

Despite decades of concern and policy guidance, the health burden attributable to indoor mould remains largely unquantified. Multiple reviews and meta-analyses have explored the relationship between indoor mould, dampness and respiratory health across an international set of studies^1–3^. A meta-analysis assessing indicators of indoor dampness and mould exposure found strong evidence of a causal relationship with asthma onset^1^. This is consistent with previous meta-analytic findings that associate building dampness and mould with a 30% to 80% increase in respiratory outcomes, reinforcing the need for preventative interventions. ^2^.

The World Health Organization’s (WHO) 2009 Guidelines on Indoor Air Quality solidified the notion that exposure to damp and mould conditions detrimentally affects respiratory symptoms and diseases such as asthma and allergies, while also posing further threats to the immunological system^4^. The guidance emphasises that indoor mould can exacerbate existing conditions like asthma and chronic obstructive pulmonary disease (COPD), and likely contributes to increased incidence of lower respiratory tract infections (LRTIs).

Yet, while the *association* is widely accepted, efforts to quantify the population health and economic burden remain scarce. Without such estimates, it is difficult to prioritise, fund, or evaluate mould-related interventions- whether they involve moisture control measures, ventilation upgrades, or broader housing policy reform ^5,6^. Justifying and prioritizing these interventions, especially on a cost-effectiveness basis, necessitates a comprehensive assessment of the health and economic impacts that such measures would bring about.

This study quantifies the health benefits and economic impacts of completely eliminating indoor mould. Specifically, it focuses on three primary respiratory health pathways: 1) the reduction in asthma incidence, 2) the attenuation of COPD severity, and 3) the decrease in the Lower LRTIs incidence. Using an open-cohort model incorporating births in future years, we estimate the health benefits, healthcare cost savings, health equity impacts, and productivity gains of mould eradication in the Australian population from 2021 onward. While this study focuses on Australia, the health risks posed by mould are universal, making these findings relevant to other countries and policy contexts.

The analysis is framed within a hypothetical scenario wherein indoor mould is completely eliminated through what could be imagined as a ’magic wand’ intervention, aiming to measure the uppermost potential impact of such a public health advancement. There is considerable uncertainty that the rigour of trying to estimate the burden from mould in models such as these lays bare; another objective of this study, therefore, is to identify key research priorities needed to strengthen our understanding of mould- related health effects.

## 2. METHODS

The study had three main methods steps. First, the intervention and Business-As-Usual (BAU) scenarios were conceptually defined. Second, input parameters for a proportional multistate lifetable (PMSLT) model were specified, incorporating population demographics, disease epidemiology, mould prevalence, causal links between mould and health outcomes, as well as health expenditure and income data.

Third, analyses were conducted using an open cohort model that included projected births, based on 2021 Australian Bureau of Statistics data, over 10-, 20-, and 40-year time horizons.

### 2.1 Conceptualisation of BAU and the intervention

The intervention is defined as the complete and permanent removal of indoor mould, contrasted with a Business-As-Usual (BAU) scenario in which existing mould prevalence remains constant over time. Health impacts were modelled via three causal pathways: reductions in asthma incidence, improvements in COPD severity, and decreases in the incidence of lower respiratory tract infections (LTRIs). We assumed that these health effects would occur immediately following intervention implementation, with no time lag. (Figure 1)

**Figure 1.**
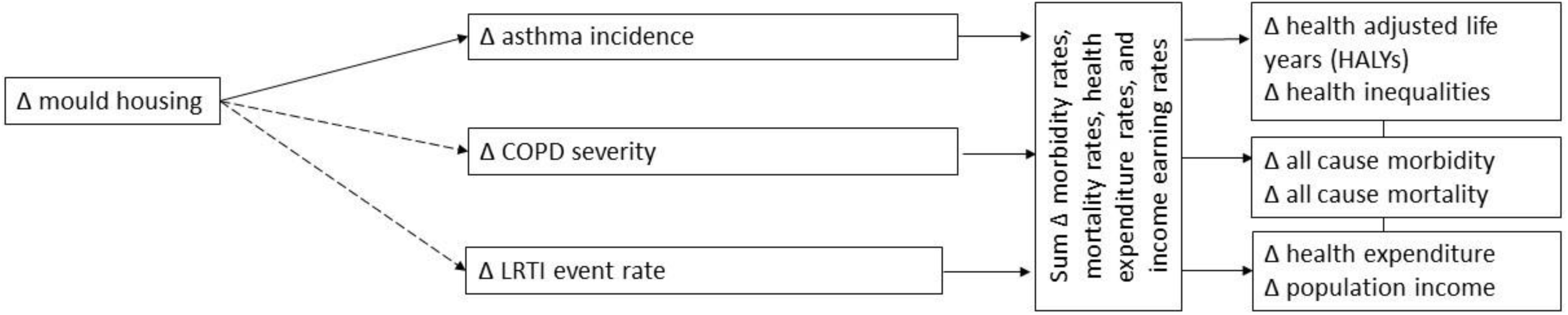
Conceptual diagram for the relationship between indoor mould and health outcomes Solid-line arrows to diseases are those where the evidence on the magnitude of indoor mould to disease to be ‘strong’. Dashed ‘moderate’ arrows are where we accept the disease is causally associated, but the magnitude of the association is poorly ascertained in previous research requiring us to ‘cross-walk’ and use other methods to specify the effect size, and COPD–- chronic obstructive pulmonary disease.

### 2.2 Model specification

We used a proportional multistate lifetable (PMSLT) model to quantify the health and economic effects of the intervention on the Australian population, starting from the year 2021. The PMSLT operates in two sequential stages: first, it models the effect of reduced mould prevalence on the incidence of selected diseases; second, it estimates the resulting changes in morbidity and mortality to derive impacts on health-adjusted life years (HALYs), healthcare costs, and income. A detailed explanation of the PMSLT framework can be found elsewhere ^7^. Key input parameters are summarised in Table 1 and described in the following sections.

**Table 1.** Input parameters for PMSLT model.

| Parameter | Data Source | Comments/ notes/ model and data assumptions |  |  |  |  |  |
| --- | --- | --- | --- | --- | --- | --- | --- |
| *Demography* |  |  |  |  |  |  |  |
| Distribution of individuals by sex, age and SEIFA quintile | ABS (2020) | Data on distribution of individual by SEIFA quintile was obtained from ABS. |  |  |  |  |  |
| Exposure (indoor mould) |  |  |  |  |  |  |  |
| Proportion living in indoor mould | AHCD (2020) | Age category | SEIFA | mean prevalence of mild mould | Standard deviation of mild mould | Mean Prevalence of severe mould | Standard Deviation of severe mould |
|  |  | 0-17 | SES1 | 0.1896 | 0.0086 | 0.1495 | 0.0079 |
|  |  | 0-17 | SES2 | 0.1688 | 0.008 | 0.1265 | 0.0072 |
|  |  | 0-17 | SES3 | 0.1814 | 0.0084 | 0.1216 | 0.0072 |
|  |  | 0-17 | SES4 | 0.1673 | 0.0078 | 0.1142 | 0.0067 |
|  |  | 0-17 | SES5 | 0.1981 | 0.0085 | 0.1096 | 0.0067 |
|  |  | 18-29 | SES1 | 0.18 | 0.0094 | 0.1281 | 0.0082 |
|  |  | 18-29 | SES2 | 0.1592 | 0.0085 | 0.1077 | 0.0072 |
|  |  | 18-29 | SES3 | 0.1711 | 0.0092 | 0.1036 | 0.0073 |
|  |  | 18-29 | SES4 | 0.1574 | 0.0085 | 0.097 | 0.0068 |
|  |  | 18-29 | SES5 | 0.1868 | 0.0093 | 0.0932 | 0.0066 |
|  |  | 30-49 | SES1 | 0.1896 | 0.0086 | 0.1495 | 0.0079 |
|  |  | 30-49 | SES2 | 0.1688 | 0.008 | 0.1265 | 0.0072 |
|  |  | 30-49 | SES3 | 0.1814 | 0.0084 | 0.1216 | 0.0072 |
|  |  | 30-49 | SES4 | 0.1673 | 0.0078 | 0.1142 | 0.0067 |
|  |  | 30-49 | SES5 | 0.1981 | 0.0085 | 0.1096 | 0.0067 |
|  |  | 50-64 | SES1 | 0.1471 | 0.0089 | 0.1113 | 0.0081 |
|  |  | 50-64 | SES2 | 0.129 | 0.0084 | 0.0927 | 0.0073 |
|  |  | 50-64 | SES3 | 0.1389 | 0.0089 | 0.0894 | 0.0072 |
|  |  | 50-64 | SES4 | 0.1272 | 0.0084 | 0.0833 | 0.0069 |
|  |  | 50-64 | SES5 | 0.1519 | 0.0095 | 0.0806 | 0.0068 |
|  |  | 65+ | SES1 | 0.0932 | 0.0108 | 0.093 | 0.0113 |
|  |  | 65+ | SES2 | 0.0806 | 0.0096 | 0.0765 | 0.0098 |
|  |  | 65+ | SES3 | 0.0872 | 0.0103 | 0.074 | 0.0095 |
|  |  | 65+ | SES4 | 0.0792 | 0.0096 | 0.0685 | 0.009 |
|  |  | 65+ | SES5 | 0.0957 | 0.0112 | 0.067 | 0.0089 |
| Time exposed (Y) | Assumed | We assumed that indoor mould impacts the prevalence of the diseases through individual's total exposure history. This time exposed was then used with the intervention effect size |  |  |  |  |  |
| *BAU – epidemiological parameters* |  |  |  |  |  |  |  |

| Parameter | Data Source | Comments/ notes/ model and data assumptions |
| --- | --- | --- |
| All-cause mortality rates | GBD | Data on all-cause mortality rates by sex and age group for 2019 were obtained from the Global Burden of disease results tool.<sup>21</sup><br>*Data on SEIFA rate ratio differences in all-cause mortality was obtained from the Australian BDS, to specify mortality RRs for disaggregation by SEIFA within our heterogeneity module (see Methods text).* |
| All-cause morbidity rates | GBD | Data on years of life lived with disability (YLD) were obtained from the Global Burden of Disease study for each sex and age group in 2019. No time trend was allowed. *Data on SEIFA differences was taken from the Australian BDS, to specific morbidity RRs for disaggregation by SEIFA.* |
| Disease specific incidence, prevalence, and case fatality rates | GBD | <u>Disease codes:</u><br>Asthma: GBD code 515; ICD codes J45-J46.0, Z82.5<br>COPD: GBD code 509; ICD-10 codes J41-J42.4, J43-J44.9<br>LRTI: GBD code 322; ICD-10 code(s) A48.1, A70, B96.0-B96.1, B97.21, B97.4-B97.6, J09-J18.2, J18.8-J18.9, J19.6-J22.9, J85.1, J91.0, P23-P23.9, U04-U04.9, Z25.1<br><u>Age by SEIFA:</u><br>To disaggregate by SEIFA, we used the following process:<br>1) We sourced mortality and prevalence rates by SEIFA from the 2018 Australian BDS. We then fitted a simple log-link regression model by sex separately for mortality (Poisson error) and prevalence (binomial error) with main effects for age (five-year age categories) |
|  |  | and SEIFA (quintiles coded as integer continuous) and an interaction term of age (aggregated categories 0-14, 15-24, 25-44, 45-64, 65-74, 75-84 and 85+) with integer SEIFA.<br>2) If for most age-groups, the SEIFA gradient in mortality was between 0.8 to 1.25 of that for prevalence:<br>a) We assumed no SEIFA differences in case fatality rates (CFR)<br>b) We assumed the RRs for incidence were those observed for mortality.<br>3) If for most age-groups, the SEIFA gradient in mortality was less than 0.8 or greater than 1.25 of that for prevalence:<br>a) We assumed SEIFA differences in case fatality rates (CFR), given by the ratio of mortality RRs to prevalence RRs.<br>b) The incidence RR was [Prevalence RR] * [CFR RR].<br>*Uncertainty:* SD was calculated by dividing the log of length of uncertainty interval (upper limit- lower limit) for disease specific incidence, prevalence, and case fatality rates by 3.92.<br><u>Annual Percentage Change (APC):</u><br>All-cause mortality rates by sex and 5-year age groups for Australia were obtained from GBD. For each sex, annual percentage changes (APCs) were estimated using Poisson regression. |
|  |  | Interaction terms between age groups and year in separate models by sex, allowing for age variations in all-cause mortality rates.<br>$\text{Ln}[\text{all-cause mortality rate}] = \text{intercept} + B1.\text{Age}[5\text{yr cat}] + B2.\text{Year} + B3.\text{Age}[1-19].\text{Year} + B4.\text{Age}[20-34].\text{Year} + B5.\text{Age}[35-49].\text{Year} + B6.\text{Age}[50-79].\text{Year} + B7.\text{Age}[80+].\text{Year}$<br>Where: B1, B2, ..., B7 are the model coefficients, with B1 actually being a vector of coefficients for all five-year age categories. Age[5yr cat] is the categorical variable for 5-year age groups. Year is calendar year as a continuous variable. Age[1-19], Age[20-34], ..., Age[80+] are dummy variables for these age groups, used in interactions with calendar year. The coefficients B2 to B7 were then converted into APCs in mortality by age, and served as inputs to the model. |
| Disease specific morbidity | IHME/GBD | The sex and age specific disability rates were calculated as disease's YLD obtained from GBD <sup>21</sup> divided by the prevalent cases.<br>The same disability rate was assumed by SEIFA (i.e., those with disease are assumed to have same severity distribution across SEIFA). |
| *Health expenditure* |  | Health expenditure per disease group data from AIHW health expenditures, stratified into three distinct disease phases (e.g., incident year, prevalent year and last year of life) (further description is available elsewhere <sup>18</sup>)<br>*Uncertainty: +/- 10% SD* |
| *Income loss* |  | Income loss per disease group data from NZ (given paucity of Australia specific income loss data), stratified into three distinct disease phases (e.g., incident year, prevalent year and last year of life) (further description is available elsewhere <sup>18</sup>) |

| Parameter | Data Source | Comments/ notes/ model and data assumptions |
| --- | --- | --- |
|  |  | *Uncertainty: +/- 10% SD* |

#### 2.2.1 Mould prevalence

Indoor mould is complex to define, operationalize and measure. For simplicity, we defined indoor mould as “houses with mould growth (visible mould or odour) in the living spaces (lounge, bedrooms) when in use by occupants.” ^4^ Not all the input data and studies used in this paper accord exactly with this definition, but it acts as our benchmark.

To specify exposure to indoor mould in Australia, we used data from the Australian Rental Housing Conditions Data infrastructure (ARHCD) (n=15,004), which provides information on the household demographics, housing conditions, and dwelling quality, collected from representative samples of public and private tenants across Australian states and Territories between July and August 2020^8^. We linked the ARHCD with Socio- Economic Indexes for Areas (SEIFA) data provided by the Australian Bureau of Statistics (Australian Bureau of Statistics, 2016), leading to a final sample of 13,882 observations. Indoor mould was assessed by two questions- “Are you experiencing any of the following problems with your current home? – Mould”, and “Do any of these need urgent repair- Mould”. Severe mould was indicated by participants answering affirmably to both questions (11.0%) and moderate mould by answering yes to the first question only (16.5%). The prevalence of mould by age group (0-17, 18-29, 30-49, 50-64, and 65+ years) cross-classified by SEIFA (quintiles 1 to 5) is shown in Figure 2: mould was less common among older people, and severe mould was more common for the most deprived compared to least deprived respondents (odds ratio 1.43).

**Figure 2:**
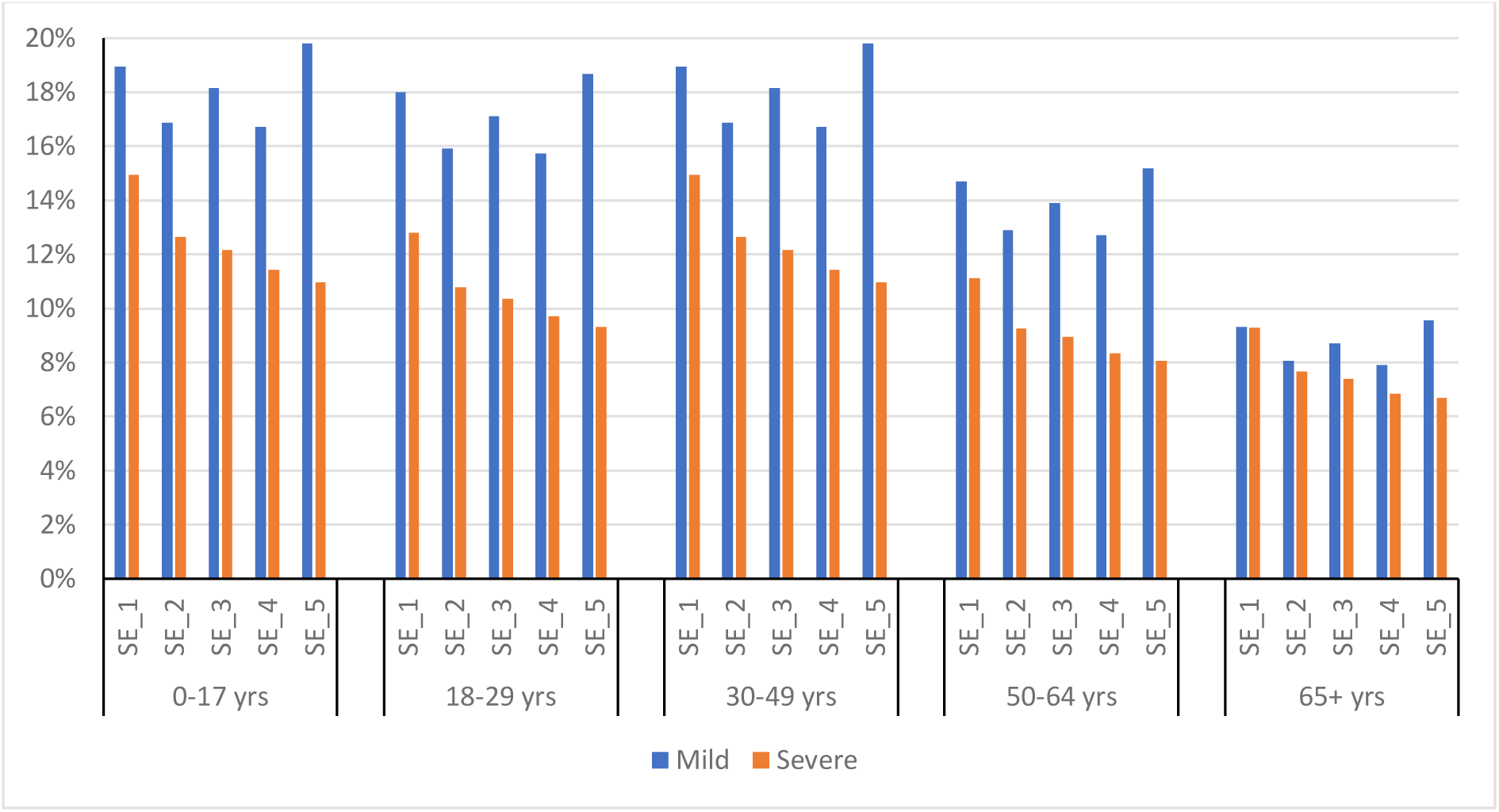
Predicted prevalence of moderate and severe mould from multinomial logistic model on Australian Rental Housing Conditions Data, by age group and socioeconomic index (SEIFA; SE_1 most deprived quintile, SE_5 least deprived quintile)

#### 2.2.2 Causal effect of indoor mould on disease incidence rates or severity

To estimate of the effect sizes of indoor mould on respiratory diseases, a systematic literature review was conducted, as described in Appendix 1. A priori, in the absence of randomized trials, the ‘best’ studies would be cohort studies that measured mould objectively at the outset, then followed up participants for (preferably) objectively determined incidence of new respiratory disease among healthy or random samples of the population (or severity of disease among representative samples of those with pre-existing disease), and fully adjusted for potential confounders of any association of mould with respiratory disease incidence or severity. Unfortunately, no such ideal study existed. Furthermore, for chronic obstructive respiratory disease and lower tract respiratory infections, most studies were cross-sectional, with self-reported ascertainment of both mould and symptomology (likely biasing the studies to report a positive association due to dependent measurement error of exposure and outcome assessment). Accordingly, we critically appraised the studies to identify the ‘best’ studies (i.e. those likely to have the least bias) and used those to quantify effect sizes as described below.

##### 2.2.2.1 Asthma incidence

Three studies ^9, 10 11^ followed up cohorts for asthma occurrence. Further details on exposure and outcome assessments are provided in Appendix 1.

To derive estimates of the asthma incidence rate ratio for moderate and severe mould with the relative frequency in Australia (above), we ran a ln-linear meta-regression model across the three studies with each study’s severity groupings coded as the midpoint on a cumulative distribution. For example, if 10% of people severe mould, and 15% of people in a given study have moderate mould, the former group was coded as 0.95 (as 0.95 is the midpoint of the top 10% of mould that was severe) and the moderate group was coded as 0.825 (the midpoint on the cumulative distribution for the 15% of people with moderate mould). The no mould group was located at 0.375, the midpoint of the 75% of people with no mould. The intercept was fixed at zero. Each study’s mould RR was weighted by the inverse of its variance. We then used slope for the cumulative rank of mould to ’read off’ the RR values for our categorization of mould, giving RRs of 1.48 for mild/moderate mould compared to nil mould (95% CI: 1.36, 2.26) and 1.98 for severe mould (1.61 to 2.24).

##### 2.2.2.2 Chronic obstructive pulmonary disease

A meta-analysis by Fisk et al (2010)^12^, the included individual studies, and further studies including those post Fisk et al^12^, were retrieved and detailed as shown in Appendix 1. No adult cohort studies of either incidence or severity of symptoms fitted our conceptualisation and eligibility criteria. Only one cross-sectional study, Haverinen et al (2001)^13^, had an objective assessment of exposure: dampness (not mould per se) in three levels of severity (nil (66.2% of 624 houses), moderate (18.5%) and severe (15.3%)) conducted by independent inspectors (meaning that at least dampness measurement was independent of participants’ self-reported symptoms) and separately self-reported by participants. The outcome measures included a battery of 23 self-report questions of symptoms in the last 12 months (e.g. bronchitis, cough, wheeze) that were grouped into five domains. We used the ‘lower respiratory symptoms’ group (including cough with phlegm, cough without phlegm, nocturnal cough, dyspnoea and wheezing) as a proxy for exacerbation of chronic obstructive pulmonary disease symptoms of severity. The multivariable adjusted risk ratio for these symptoms among people in moderately damp housing compared to non-damp housing was 1.04 (95% CI 0.83 to 1.30) and in severely damp housing was 1.33 (95% CI 1.07 to 1.65). To estimate the risk ratios given our distribution of mould, assuming a linear dose-response within the moderate and severe categories, we ran an intercept-free linear regression by the midpoint above nil (i.e., no mould) weighted using inverse variance of each category.

The estimates were 1.10 (95% CI 0.88 to 1.37) for our moderate category and 1.28 (95% UI 1.03 to 1.59) for our severe category, each compared to nil mould. These risk ratios were assumed to apply to COPD disease severity in the PMSLT. We truncated all risk ratio draws at 1, under the assumption that mould is not protective for any of the included diseases.

##### 2.2.2.3 Lower respiratory tract infection

As with COPD above, Haverinen et al (2001)^13^ was the best study to use for a ‘respiratory infections’ grouping of outcomes that we used as a proxy for LRTI occurrence (acknowledging some of the questions were more specific to upper respiratory tract infections). Using the same methodology as above for COPD, the derived risk ratios applied to LRTI event rates were 1.15 (95% CI 0.88 to 1.51) for our moderate category and 1.46 (95% CI 1.13 to 1.90) for our severe category . All risk ratio draws were left-truncated at 1 under the assumption the mould exposure is not protective of health.

Percentage reduction in disease incidence and morbidity from eradicating indoor mould For asthma and LRTI, for each age by SEIFA group, a population impact fraction (PIF) was calculated :

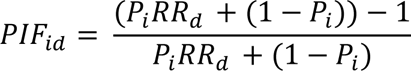

where: *I* indexes each age-by-SEIFA group and *d* refers to each disease; *P*_*i*_ is the percentage of the population exposed to indoor mould in group *i*; and *RR*_*d*_ is the relative risk comparing those exposed to indoor mould with those unexposed for disease *d*. *P*_*i*_ is assumed to remain constant over time in the BAU scenario, resulting in time-invariant PIFs. A zero-time lag was also assumed, whereby changes in mould exposure immediately affect disease rates.

The above PIF is usually for disease incidence or event rates. We used the same formula for percentage reduction in COPD morbidity.

#### 2.2.3 Demographics and disease epidemiology

Baseline population counts by sex, age, and SEIFA were obtained from 2021 Australian Bureau of Statistics (ABS) data. Future population growth was modelled using the ABS’s moderate fertility projection scenario. Migration was not included.

Forecasts of prevalence, mortality and morbidity for each disease were obtained by assuming log-linear trends on GBD data from 1991 to 2019. Incidence, remission, and case fatality rates were estimated similarly, then calibrated to attain the prevalence, mortality and morbidity forecasts. A log-linear trend on calibrated rates is applied until 2034 via annual percentage change, and beyond 2034 the rates are kept constant.

Baseline disease prevalence and mortality rates by sex and age were disaggregated by SEIFA quintile using rate ratios (RRs) from the Australian Burden of Disease Study, following an approach consistent with our previously published heterogeneity module ^14,15^. SEIFA-specific RRs were derived from log-link regression models applied separately by sex: Poisson models for mortality and binomial models for prevalence.

Each model included five-year age groups and SEIFA quintiles, with interaction terms between SEIFA and age to account for age-specific socioeconomic gradients.

From these modelled differences, SEIFA-specific case fatality rate ratios were derived by dividing mortality RRs by prevalence RRs, using the formula *[RR of mortality rates by SEIFA] / [RR of prevalence by SEFIA].* Disease incidence RRs were then calculated as the product of prevalence RRs and case fatality RRs, using *[RR of prevalence by SEIFA]*[RR of case fatality rate by SEFIA]*. All-cause mortality and morbidity (measured as years lived with disability) were modelled using a similar approach based on 2019 AIHW data.

### 2.3 Analyses

The proportional multistate lifetable model was executed for both the Business-As-Usual and intervention scenarios, with the intervention defined as the hypothetical elimination of indoor mould. In the intervention scenario, disease incidence and event rates were adjusted using the population impact fractions described above. To incorporate uncertainty, each model was run 2,000 times with probabilistic sampling across all input parameters (see Table 1). Analyses were conducted over three time horizons: 10, 20, and 40 years from a 2021 baseline.The model generated the following key outcomes: 1) Health-adjusted life years (HALYs) gained from mould eradication, stratified by disease and SEIFA quintile; 2) Healthcare cost savings, calculated by applying AIHW disease-specific expenditure estimates ^16^; 3) Income gains, estimated using purchasing power parity–adjusted New Zealand data on income differentials between individuals with and without disease ^17^, mapped to disease states and time periods in line with expenditure patterns (see Table 1; further description is available elsewhere ^18^). All outcomes were calculated using a 3% annual discount rate, with undiscounted results provided in Appendix 2.

To assess the intervention’s impact on health inequality, we calculated the age standardised ratio of HALYs gained between the most and least deprived socioeconomic groups. To do so we divided the HALYs gained by each age group, within each year and strata, by the corresponding person years, then weighted the result using the WHO standard population weights.

To assess the impact of input uncertainty on model outcomes, we conducted univariate sensitivity analyses. For each key parameter, we evaluated model outputs using its 2.5th and 97.5th percentile values, while holding all other inputs at their expected values.

This approach allowed us to isolate the contribution of individual inputs to the overall uncertainty in HALYs gained.

Results are presented as a tornado plot, where the central line represents the median HALY estimate and the length of each bar reflects the range of variation due to each parameter.

All analyses were carried out in Python (version 3.9.5, Delaware, USA).

## 3. RESULTS

Table 2 shows the HALYs gained, health expenditure savings, and income gains for eradication of indoor mould from 2021. Discounted at 3% per annum, eradicating mould leads to 109,000 (95% uncertainty interval (UI): 50,100 to 191,000) total HALYS gained in the next 20 years. Expressed alternatively, that is 4,190 (95% UI 1920 to 7350) HALYs gained per million people alive in 2021 in the next 20 years (or about 1.5 health days per person). Disaggregated by diseases, most HALY gains was contributed by reductions in asthma incidence (median HALYs gain 46,900, 95% UI 22,600 to 98,100) and COPD morbidity rates (median HALYs gain 46,900, 95% UI: 3,530 to 116,000). The population weighted percentage reduction in LRTI and Asthma was 7% and 15%, respectively and the percentage reduction in COPD morbidity was 4%.

**Table 2:** Future HALYs gained, health expenditure saved, and income gained for eradication of indoor mould compared to BAU, discounted at 3%.

|  | 2021-2031 | 2031-2041 | 2041-2061 | 2021-2041 | 2021-2061 |
| --- | --- | --- | --- | --- | --- |
| Health adjusted life years gained compared with business as usual |  |  |  |  |  |
| Total HALYs gained (millions) | 44,600<br>(18,500 to<br>81,500) | 64,200 (31,000 to<br>111,000) | 125,000 (61,700<br>to 212,000) | 109,000 (50,100 to<br>191,000) | 233,000 (112,000 to<br>402,000) |
| By quintile of SEIFA |  |  |  |  |  |
| Most deprived quintile | 11,300 (5,030<br>to 20,100) | 16,300 (8,320 to<br>27,500) | 30,500 (15,900 to<br>51,000) | 27,800 (13,600 to<br>47,300) | 58,500 (29,500 to<br>97,900) |
| 2<sup>nd</sup> most deprived quintile | 9,210 (3,840<br>to 16,700) | 13,100 (6,390 to<br>22,400) | 24,800 (12,500 to<br>41,900) | 22,300 (10,300 to<br>39,000) | 47,200 (23,100 to<br>80,700) |
| 3<sup>rd</sup> most deprived quintile | 8,810 (3,690<br>to 16,100) | 12,800 (6,150 to<br>22,100) | 25,000 (12,400 to<br>42,500) | 21,600 (9,930 to<br>37,900) | 46,500 (22,400 to<br>80,400) |
| 4<sup>th</sup> most deprived quintile | 7,640 (3,150<br>to 14,000) | 11,100 (5,260 to<br>19,400) | 22,500 (10,900 to<br>38,700) | 18,800 (8,540 to<br>33,400) | 41,400 (19,700 to<br>71,800) |
| Least deprived quintile | 7,550 (2,810<br>to 14,600) | 10,600 (4,710 to<br>19,500) | 21,600 (10,000 to<br>38,800) | 18,300 (7,630 to<br>34,100) | 39,900 (17,500 to<br>73,000) |
| Ratio for most vs least deprived ‡ | 1.64 (1.40 to 2.00) | 1.68 (1.41 to 2.01) | 1.44 (1.20 to 1.82) | 1.66 (1.40 to 2.01) | 1.54 (1.29 to 1.90) |
| By sex |  |  |  |  |  |
| Female | 23,400 (9,390 to 43,300) | 35,300 (16,500 to 61,600) | 71,200 (34,400 to 122,000) | 58,700 (26,300 to 105,000) | 130,000 (60,800 to 227,000) |
| Male | 21,200 (9,220 to 38,200) | 28,700 (14,300 to 48,400) | 53,300 (27,000 to 90,000) | 49,800 (23,700 to 86,400) | 104,000 (50,900 to 175,000) |
| By disease |  |  |  |  |  |
| Asthma incidence | 16,800 (8,030 to 34,700) | 30,200 (14,500 to 63,400) | 55,800 (26,800 to 116,000) | 46,900 (22,600 to 98,100) | 102,000 (49,400 to 216,000) |
| COPD severity | 23,000 (1,750 to 55,800) | 23,900 (1,780 to 60,100) | 42,100 (3,140 to 108,000) | 46,900 (3,530 to 116,000) | 89,000 (6,620 to 223,000) |
| LRTI event rate | 3,330 (707 to 7,450) | 7,190 (1,520 to 16,200) | 21,100 (4,410 to 48,600) | 10,500 (2,230 to 23,800) | 31,700 (6,670 to 71,900) |
| Health expenditure change (\$millions in 2021 AUD) compared with business as usual | | | | | |
| Total | -1,450 (-2,630 to -611) | -1,370 (-2,380 to -641) | -1,930 (-3,390 to -925) | -2,820 (-4,990 to -1,250) | -4,750 (-8,290 to -2,190) |

| Income gain (\$millions in 2021 AUD) compared with business-as-usual | | | | | |
| --- | --- | --- | --- | --- | --- |
| Income gain | 1,770 (659 to 3,420) | 2,420 (1,100 to 4,620) | 4,210 (2,090 to 8,240) | 4,210 (1,760 to 7,910) | 8,450 (3,890 to 16,100) |

‡Age standardized to WHO World population divided by person year

Greater HALY gains were observed in lower socioeconomic groups. The age standardised ratio of HALYs gained between the most and least deprived groups in the next 20 years is 1.66 (95 UI 1.40 to 2.01). That is, removing mould would generate 1.66 times greater health gain for the lowest SES groups who have highest disease rates.

Health expenditure was estimated to be cumulatively reduced by AUD2,820 million (95% UI 1,250 to 4,990, 3% discount rate) in the next 20 years compared to the BAU scenario. That is US$ 78.0 million per million people (95% UI34.6 to 138). Expressed alternatively, it is 0.5% to 2.1% of all health spending in Australia in a year or 0.7% to 2.8% of government health spending in a year.

Population income was predicted to increase by AUD$4,210 million (95% UI 1,760 to 7,910, 3% discount rate) due to increased productivity, or US$ 116 million (95% UI 48.7 to 219), Expressed alternatively, this income gain is equivalent to 0.08% to 0.36% of Australia’s GDP in 2021.

Figure 3 presents a tornado plot illustrating the extent to which variation in the HALYs gained over the first 20 years is attributable to uncertainty in individual model inputs. The top bar represents the total uncertainty range from the full probabilistic (Monte Carlo) analysis. Each subsequent bar shows the impact of varying a single input parameter across its 2.5th and 97.5th percentile values, while holding all other parameters at their median estimates. Uncertainty in the association between indoor mould and COPD severity accounted for the largest proportion of overall variation in HALY estimates. Specifically, when varying the rate ratios for both mild and severe mould exposure on COPD severity across their 95% uncertainty interval, HALYs gained ranged from 62,400 to 176,000. In contrast, variation in other key input parameters contributed substantially less to the total uncertainty in HALY outcomes.

**Figure 3:**
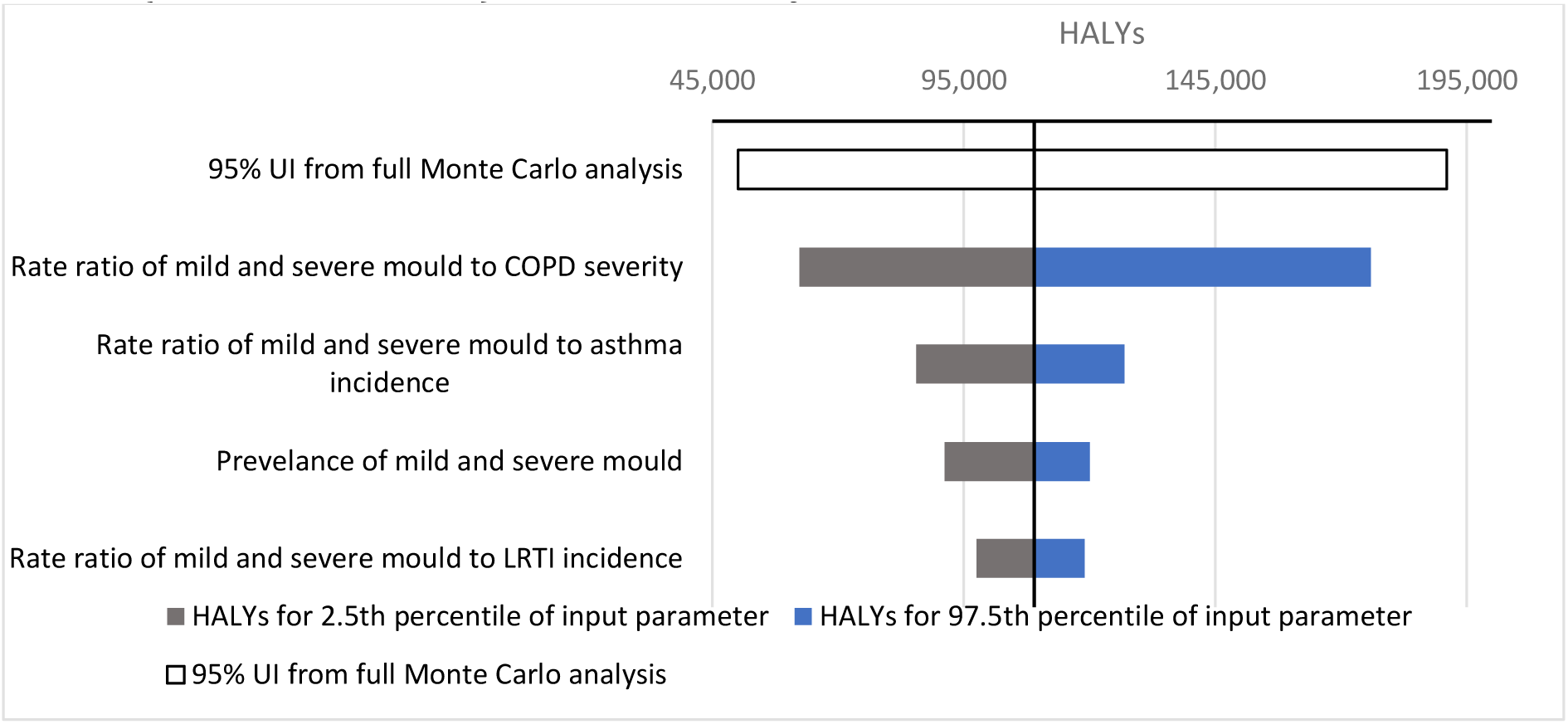
Tornado plots showing input parameters ranked in terms of their 2.5th and 97.5th percentile value impacts on total HALYs (3% discount rate) for the first 20 years †The vertical axis is at the HALYs value when all input parameters are at their median value. The bars show the variation in HALYs for the 2.5th and 97.5th percentile values of the input parameter.

## 4. DISCUSSION

This study models the health and economic impacts of a hypothetical ‘magic wand’ intervention of eradicating indoor mould, for a case study of the nearly 28% of Australian households at risk^4^. Through reductions in asthma incidence, COPD severity, and LTRI incidence, this intervention may lead to 109,000 (95% UI 50,100 to 191,000) HALYs gained in the next 20 years, or 4190 (95% UI 1920 to 7350) HALYs gained per million people alive in 2021 in the next 20 years (about 1.5 health days per person).

The potential health gain in 20 years horizon is two times higher compared to that achieved from eradicating cold housing in Australia; estimated to be 50,600 HALYs (24,600 to 106,000) by using the same methods but with a closed-cohort ^19^.

We estimated the potential direct health expenditure savings by removing indoor mould to be AUD$ 2.82 billion from 2021 to 2041 or US$ 78.0 million per million people (95% UI34.6 to 138). This saving is equivalent to 0.5% to 2.1% of all health spending in Australia in a year or 0.7% to 2.8% of government health spending in a year. From another perspective, AUD$2.82 billion is about two thirds of an estimated $4.5 billion of expenditure in the Australian health system was for respiratory conditions in 2020- 21 ^20^. The other economic impact estimated in this study was an increase of income in the next 20 years by US$ 116 million (95% UI 48.7 to 219), equivalent to 0.08% to 0.36% of Australia’s GDP in 2021. It is important that preventive programs in health are examined for their potential to reduce (or delay) health system expenditure, and to increase income productivity so as to assist society manage increasingly aging populations. In this light, and not quantified in this paper, preventative interventions can not only increase workforce income productivity among the ‘<65-year-old working age population’, but also allow people who wish to the ability to work beyond the age of 65 years if morbidity is reduced. This is an important area to explore more in future work.

This study provides an upper-bound estimate of potential benefit—not as a policy prescription, but as a starting point. Quantification is a prerequisite for action: unless we estimate how much harm indoor mould causes, we cannot meaningfully prioritise, fund, or evaluate strategies to address it. Future work can build on this foundation by assessing the cost-effectiveness of real-world interventions. Mould growth can be remediated through low-cost interventions leading to cost savings^5,6^. There are also more structural interventions, such as R installing mould resistant building materials either at initial construction or as part of (likely more expensive) retrofitting. Other interventions include using vapour barriers, increasing ventilation by keeping the windows open, not using living areas for laundry drying and using humidifiers if possible^5,6^. Heavily infested housing stock require professional remediation and repair (halting further damage to interior walls and stopping any microbial growth). There is a need to evaluate many of these interventions, separately and as packages, to estimate the optimal or most cost-effective interventions.

A strength of this study is its transparent handling of uncertainty. We conducted the modelling despite clear gaps in the evidence base, particularly regarding the causal relationship between mould exposure and disease outcomes. However, we argue that modelling should not wait for perfect data—rather, it can serve as a tool for identifying critical knowledge gaps and directing future empirical work. In that spirit, our findings suggest three research priorities. First, a major source of uncertainty lies in the causal relationship between indoor mould exposure and respiratory health—particularly for conditions such as COPD and LTRIs. While guidelines such as the WHO Housing and Health Guidelines ^4^ point to a likely association, robust and precise effect estimates are still lacking. This study contributes by not only quantifying the extent of that uncertainty but also highlighting the key drivers, thereby identifying critical evidence gaps. One clear priority for future research is to better quantify the impact of mould exposure on COPD severity. Second, accurate measurement of population exposure to indoor mould, including the prevalence, intensity, and distribution across socioeconomic groups, is needed to improve model input. More reliable exposure data would substantially improve estimates of the potential health benefits from mitigation strategies. Finally, there is an urgent need for well-designed intervention studies— ideally randomised trials—that evaluate the real-world impact of mould remediation strategies (such as improved ventilation) on objectively measured health outcomes.

Strengthening this evidence base will be essential for guiding effective public health action.

Despite these limitations, this study demonstrates that eradicating indoor mould has the potential to deliver substantial health and economic benefits for the Australian population. The gains—measured in HALYs, reduced healthcare costs, and increased income—highlight the value of addressing indoor environmental risks as a public health priority. Importantly, such interventions may also contribute to reducing socioeconomic health inequalities, given the higher burden of mould exposure among disadvantaged groups. While this modelling represents a hypothetical ‘best-case’ scenario, it underscores the urgent need for investment in research to better quantify causal relationships, improve exposure assessment, and rigorously evaluate the real-world effectiveness and cost-effectiveness of mould remediation strategies. Strengthening this evidence base will be critical for guiding equitable and impactful public health interventions in housing.

## Data Availability

This study used de-identified data from the Australian Housing Conditions Dataset 2022 (AHCD22), published through the Australian Data Archive (DOI: 10.26193/SLCU9J).

https://dataverse.ada.edu.au/dataset.xhtml?persistentId=doi:10.26193/SLCU9J

