## Supplementary material for "Quantifying Health Gains and Health System Expenditure Impacts of Eliminating Indoor Mould in Homes"

#### Table of Contents

Appendix 1. Literature review on association between mould and asthma, COPD, and LTRI

1.1 Search strategy

1.2 Eligibility criteria

1.3 Characteristics of included studies

Appendix 2. Undiscounted results

#### Appendix 1: Review of literature on mould and respiratory health

Epidemiological studies provide evidence to link dampness and mould exposures to human health effects; however, in interpreting these studies, consideration must be given to their inherent strengths and weaknesses. Errors such as studying only samples instead of entire populations and bias related to sampling, measurement and confounding can distort risk estimates. In addition, the evidence from a body of studies may be distorted by publication bias, which reflects a tendency not to publish or to publish more slowly negative or equivocal findings, which may therefore not be included in the review. In interpreting epidemiological studies on dampness, the uncertainty about the causal exposures and health effects involved must also be considered.

This appendix describes our steps to overcome these challenges and how we arrived at decisions on which pairs of mould housing to disease rates to include, and with what effect sizes. For all mould to disease associations, we assume a linear dose response, with TMREL at beginning of mild/moderate mould. In most of the studies exposure-outcome relationship are derived from answer to questionnaires. Contrarily, more robust approach would be to take airborne or surface concentration of indoor pollutants (pollutants per m$^3$ of the surface area or per gram of the dust). Our evidence at best provides good proxies for the true exposure-outcome relationship and thus is subjected to misclassification and downward bias. The imminent lack of evidence about the mean risk function is due to a lack of a valid, quantitative method for assessing exposure; thus exposure-outcome relationship may have failed to demonstrate any true causal relationship. There is critical need for large-scale, high-quality studies on each exposure-outcome pair (e.g., mould and COPD)

##### 1.1 Search strategy

This review combines the conclusions of a review by the World Health Organisation (2009), covering studies up to 2007, the Institute of Medicine (2015), covering the literature up to 2013, those of a quantitative meta-analysis of findings up to 2007 on dampness, mould and respiratory health effects (Fisk, Lei-Gomez, Mendell, 2007) and a new assessment of more recent (2013 to 2023) published studies on asthma, COPD, and LTRI.

PubMed searches were performed in Oct 2021 and updated in Dec 2023 for studies published between 2013 and 2023, using the following search strategy:

| **Step 1** | **Step 2** | **Step 3** |
|---|---|---|
| **Mould** | **Household information** | **Health outcomes** |
| (mold[TIAB] OR mould[TIAB] OR damp[TIAB] OR fung*[TIAB] OR "mould spores" [TIAB] OR cladosporium[TIAB] OR penicillium[TIAB] OR aspergillus[TIAB] OR alternaria[TIAB] OR mycotoxins[TIAB] OR endotoxin[TIAB]) AND (humans[Filter]) | (Indoor[TIAB] OR Home[TIAB] OR Domestic[TIAB] OR Dwelling[TIAB] OR Residential[TIAB] OR Housing[TIAB] OR Housing[Mesh] OR House[TIAB] OR Inside[TIAB] OR Household[TIAB]) AND (humans[Filter]) | "respiratory infections"[TIAB] OR lung*[TIAB] OR pulmonary*[TIAB] OR "chronic obstructive pulmonary disease"[TIAB] OR COPD[TIAB] OR "lung function*"[TIAB] OR asthma*[TIAB] OR bronchitis[TIAB] OR Respiratory Tract Diseases [Mesh] OR respirat*[TIAB] OR respiratory[TIAB] AND (humans[Filter]) |
| ***Step 4***<br>*Step 1 AND Step 2 AND Step 3* | | |
| ***Step 5: Limit Step 4 papers.***<br>((rate*[TIAB] or inciden*[TIAB] OR prevalen* [TIAB] OR severity [TIAB] OR remission [TIAB] OR correlation [TIAB] OR association [TIAB])) OR (("all cause"[TIAB] OR "all-cause"[TIAB] OR "total"[TIAB]) AND ("mortality"[TIAB] OR "death"[TIAB] OR "survival"[TIAB])) AND (humans[Filter]) | | |

##### 1.2 Eligibility criteria

In the absences of randomized trials, the 'best' studies would be cohort studies that measured mould objectively at the outset, then followed up participants for (preferably) objectively determined incidence of new respiratory disease among healthy or random samples of the population (or severity of disease among representative samples of those with pre-existing disease), and fully adjusted for potential confounders of any association of mould with respiratory disease incidence or severity. Unfortunately, no such ideal study existed. Furthermore, for chronic obstructive respiratory disease and lower tract respiratory infections, most studies were cross-sectional, with self-reported ascertainment of both mould and symptomology (likely biasing the studies to report a positive association due to dependent measurement error of exposure and outcome assessment). Accordingly, we critically appraised the studies to identify the 'best' studies (i.e. those likely to have the least bias) and used those to quantify effect sizes.

**1.3 Characteristics of included studies**

After the first stage screening, we've identified 9 cohort studies for asthma, 6 cohort studies for COPD and LTRI (combining studies included in previous reviews). Details of the studies are presented in the tables below. Among them, three studies were included in the final estimate for asthma, and one study was cross walked to device estimate for COPD and LTRI.

**Table S1. Summary of studies on mould and asthma incidence**

| STUDY | COUNTRY | SAMPLE | EXPOSURE | OUTCOMES | CONFOUNDING FACTORS | MEASURE OF EFFECT | EFFECT ESTIMATE | Meeting criteria> |
|---|---|---|---|---|---|---|---|---|
| **Norback(2011) [Cohort study]** | Europe and USA | 6,443 | A mould score (0-4), derived by adding the number of yes answers to the four questions on indoor mould in any place in the home (indoor mould ever in the European Community Respiratory Survey (ECRHS) I or II, indoor mould in the last 12 months in ECRHS I or II). Binary exposure was used in the analyses: score >0 vs score=0. | Lung function decline (forced expiratory volume in 1 second ($FEV_1$)) measured by Spirometry. Disease progression (neither disease incidence nor disease severity per se, although one would expect a correlation). Age 20-44 years. | Age, height, BMI, length of follow-up and smoking category | Adjusted difference in decline over a 9-year period | *FEV1 (ml per year) decline:*<br>For men:<br><-0.001 (-2.50 to 2.50)<br><br>For women:<br>-0.83 (-2.76 to 1.09) | *No* |
| **Norback(2013) [Cohort study]** | Europe, USA, and AUS | 7,104 | 1. A mould score (0-4), derived by adding the number of yes answers to the four questions on indoor mould in any place in the home (indoor mould ever ECRHS I or II, indoor mould 12 months ECRHS I or II).<br><br>2. Number of rooms with mould<br><br>3. Mould location (e.g., bedroom, living room, bathroom) | New onset of asthma [excluding participants with wheeze, nocturnal shortness of breath in the last 12 months, or a history of asthma at baseline), l, determined by self-report (≥1 of either asthma attack on last 12 months, medication, and shortness of breath). Young adults (age not specified). | Age, sex, smoking and study centre | RR | *Incidence*:<br>Mould score and asthma<br>***1-2 score c.f. nil***<br>RR= 1.05 (0.82 to 1.33)<br>***3-4 score c.f. nil***<br>RR= 1.73 (1.27 to 2.37)<br>***Test for trend***<br>RR=1.12 (1.03 to 1.22)<br>Number of bedrooms and asthma<br>***1 score c.f. nil***<br>RR= 1.30 (1.02 to 1.66)<br>***2-3 score c.f. nil***<br>RR= 1.43 (0.83 to 2.48)<br>*Dose-response*<br>1.17 (1.04 to 1.32) | *Yes* |
| **Jaakkola(2010) [Cohort study]** | Finland | 1863 | Visible mould, mould odor, or any (additionally incl. moisture, and water damage) at baseline | Allergic Rhinitis. Age 1-7years | Age, sex, breastfeeding, education, single parents/guardian, smoking in pregnancy, secondhand smoking, gas cooking, pets, day-care, other exposures (mutually adjusting) | aOR (mould/odor/any) | Visible mould: 1.06, (0.51, 2.21)<br>Mould odor: 0.94 (0.36, 2.45)<br>Any exposure: 1.55 (1.10, 2.18) | No |
| **Jaakkola(2005) [Cohort study]** | Finland | 1,984 | Visible mould, mould odor, or any (additionally incl. moisture, and water damage) at baseline | Asthma. Age 1-7 years | Age, sex, breastfeeding, education, single parents/guardian, smoking in pregnancy, secondhand smoking/ETS, gas cooking, pets, day-care, | aOR (mould/odor/any) | Visible mould: 0.65 (0.24–1.72)<br>Mould odor: 2.44 (1.07–5.60)<br>Mould odor (no parental atopy): 2.56 (0.93–7.08)<br>Any exposure: 1.01 (0.66–1.54) | Yes |

| STUDY | COUNTRY | SAMPLE | EXPOSURE | OUTCOMES | CONFOUNDING FACTORS | MEASURE OF EFFECT | EFFECT ESTIMATE | Meeting criteria> |
|---|---|---|---|---|---|---|---|---|
| | | | | | other exposures (mutually adjusting) | | | |
| **Thacher(2016) [Cohort study]** | Children born 1994-95 recruited, followed for 16 years [Sweden] | 3798 | Exposure to mould or dampness during infancy<br>Mould odor—'Is there, or has there ever been, a smell of mildew in the home?'<br>Visible mould—'Has there been any visible mould in the home in the past year (prior to the date of the baseline questionnaire)?'<br>Dampness damage—'Is there, or has there ever been, any type of moisture damage (spots or similar) in the home?'<br>Severity of exposure score—The sum of mould and dampness indicators (ranging from 0 to 3). | Asthma—four or more episodes of wheeze in the last 12 months or one or more episode of wheeze in the last 12 months in combination with inhaled steroids<br><br>Rhinitis—eye or nose symptoms following exposure to allergens in the last 12 months and/or a doctor's diagnosis of allergic rhinitis. Age <16 years. | Sex, socioeconomic status, parental allergic disease, maternal smoking during pregnancy, parental smoking during infancy, maternal age, and older siblings | aOR | **Asthma**<br>No mould or dampness indicator Reference<br>1 indicator 1.16 (0.93–1.44)<br>2 indicators 1.37 (1.01–1.86)<br>3 indicators 1.73 (1.10–2.74)<br>**Rhinitis**<br>No mould or dampness indicator Reference<br>1 indicator 1.03 (0.87–1.22)<br>2 indicators 1.18 (0.92–1.52)<br>3 indicators 1.23 (0.82–1.85)<br>**Aeroallergen sensitization**<br>No mould or dampness indicator Reference<br>1 indicator 0.94 (0.77–1.13)<br>2 indicators 0.85 (0.63–1.14)<br>3 indicators 0.90 (0.55–1.48)<br>**Food allergen sensitization**<br>No mould or dampness indicator Reference<br>1 indicator 0.87 (0.69–1.09)<br>2 indicators 1.02 (0.73–1.42)<br>3 indicators 1.22 (0.72–2.07) | **Yes** |
| **Tiesler(2015) [Cohort study]** | 10-year-old children [Germany] | 1719 | Presence of visible mould or dampness at home | Sleep problems<br>Presence of any sleep problems, problems to fall asleep, problems sleeping through the night and a 24h sleep time of less than 9 h. Age 10 years | Study centre, sex, age and level of parental education | aOR | For any sleep problems 1.77, 1.21–2.60<br>Problem sleeping through the night 2.52, 1.27–5.00<br>a short sleep time 1.68, 1.09–2.61 | No |
| **Wang (2022) [Cohort study]** | EU: Norway, Sweden, Demark, Estonia | 17881 | Questionnaire based survey of mould (Q1: "Visible mold growth indoors on walls, floors or ceilings ('visible mould')" (yes/no); Q2: "Mold odor in one or several rooms (other than the cellar)" (yes/ no).<br>Visible mould: 7.6%<br>Mold odor: 3.7% | Questionnaire based survey on history of i) asthma before 10 y (yes/no) and 2) asthma for 10 y (yes/no). Age x-x years | parental education (RHINE III) and offspring's age (RHINE III) | aOR | No dampness/mold: 1<br>Only dampness/mold follow up 1: 1.09(0.92, 1.29)<br>Only dampness/mold follow up 2<br>Dampness mold both follow up | No |
| **Milanzi (2020) [Cohort study]** | Netherlands | 552 | Aged 12 to 16 years | FEV1 and FVC at ages 12 and 16 years | Adjusted for sex, log differences in ht, wt and age (between 12 y and | % difference in FEV1, | % difference in FEV1, growth/year (95% CI) | No |

| STUDY | COUNTRY | SAMPLE | EXPOSURE | OUTCOMES | CONFOUNDING FACTORS | MEASURE OF EFFECT | EFFECT ESTIMATE | Meeting criteria> |
|---|---|---|---|---|---|---|---|---|
| | | | | | 16 y), lung function (16 y), education, allergy, breastfeeding, smoking, SHS, gas cooking, NO2, birth wt, pets and resp infections. | growth/year (95% CI) | Early life vs very low: −0.77 (−1.05 to −0.49)<br>Mod. Late childhood vs very low: 0.05 (−0.19 to 0.30)<br>Mid-childhood vs very low: 0.04 (−0.37 to 0.46)<br>% difference in FVC growth/year (95% CI)<br>Early life vs very low: 0.56 (−0.83 to −0.28)<br>Mod. Late childhood vs very low: −0.02 (−0.26 to 0.23)<br>Mid-childhood vs very low: 0.23 (−0.63 to 0.18) | |
| **Lezmi (2020) [Cohort study]** | France | 2020 | 131 | Severe recurrent wheeze defined as those with persistent symptoms or requiring medication or having severe exacerbation. | Adjusted for birth weight, SHS, atopic dermatisis | aOR | 4.22 (1.25 to 18.2) | No |

**Table S2. Summary of studies on mould and COPD severity and LTRI event rate**

| STUDY | COUNTRY | SAMPLE | EXPOSURE | OUTCOMES | CONFOUNDING FACTORS | MEASURE OF EFFECT | EFFECT ESTIMATE | Meeting criteria? |
|---|---|---|---|---|---|---|---|---|
| Ekici et al. 2008 | Turkey | 9,853 | Dampness or visible mould during childhood | RI,B | Not clear | aOR | 1.9 (1.6 to 2.3) | No |
| Haverinen et al. 2001 | Finland | 1,017 | Homes were graded (by assessors) into three categories: I, II, III based on severity and amount of moisture damages. | RI,B | age, gender, smoking, pets indoors, marital status, education, parental atopy and atopic predisposi-tion. | aOR | B: 1.56 (0.96 - 2.52)<br>RI: 1.23 (1.03 - 1.45) | Yes |
| Stark et al. 2003 | USA | 499 | in-home fungal concentrations | RI: (croup, pneumonia, bronchitis, and bronchiolitis) in the first year. least one LRTI symptom in the first year of life. The symptoms were assessed every 2 months; primary caregiver was asked this: "Since we last spoke on (date given), has your child had a pneumonia, croup, bronchitis, or bronchiolitis diagnosed by a doctor?". | Adjusted for sex, presence of water, mold/mildew, born in winter, breastfeeding, and being exposed to other children | RR | RI: 1.34 (0.99 to 1.82) | No |
| Dales et al. 2010 | Canada | 357 | Endotoxin (EU/m3) | Illness episodes (stuffy nose, cough, wheeze, shortness of breath): acute respiratory illness in 2 years of life | university education and family income | *Beta coefficient* | <u>*Endotoxin (EU/m3):*</u><br>Beta: 0.01, SE: 0.00, *P*=0.0116 | No |
| Chen et al,2012 [cohort study] | Taiwan | 21248 infants followed from birth to 6 months | Visible mould on walls | Carer reported pneumonia | Gender, preterm birth, congenital cardiopulmonary diseases, use of antibiotics during pregnancy, maternal overweight before pregnancy, prenatal ETS exposure daily, maternal tobacco smoking | aOR | aOR=1.45 (95%CI 1.02-2.06) | No |
| Lu et al, 2022 [retrospective cohort study] | China | 8689 pre-schoolers | mold/damp stains,<br>mold or damp for clothing or bedding | Carer reported pneumonia | Age, sex, birth season, breastfeeding duration, parental annual income, living area, number of people in residence, parental smoking, cooking fuel, fresh air filter | aOR | <u>Pre-birth:</u><br>Mould/damp stains:<br>aOR=1.25(1.09,1.42)<br>Mould/damp clothing or bedding:<br>1.32(1.12,1.55)<br><br><u>Post birth</u><br>Mould/damp stains:<br>aOR=1.25(1.10,1.42)<br>Mould/damp clothing or bedding:<br>1.17(0.99,1.38) | No |

| STUDY | COUNTRY | SAMPLE | EXPOSURE | OUTCOMES | CONFOUNDING FACTORS | MEASURE OF EFFECT | EFFECT ESTIMATE | Meeting criteria? |
|---|---|---|---|---|---|---|---|---|
| | | | | | | | The detrimental effects also accumulate with time | |

### Appendix 2 Undiscounted results

**Table S3. Future HALYs gained, health expenditure saved, and income gained for eradication of indoor mould compared to BAU, discounted at 0%**

| | 2021-2031 | 2031-2041 | 2041-2061 | 2021-2041 | 2021-2061 |
|---|---|---|---|---|---|
| Health adjusted life years gained compared with business as usual | | | | | |
| Total HALYs gained (millions) | 52,400 (22,000 to 95,700) | 101,000 (48,500 to 173,000) | 307,000 (153,000 to 522,000) | 153,000 (71,000 to 267,000) | 461,000 (225,000 to 786,000) |
| By quintile of SEIFA | | | | | |
| Most deprived quintile | 13,400 (5,990 to 23,600) | 25,500 (13,000 to 43,000) | 75,000 (39,200 to 126,000) | 39,000 (19,200 to 66,200) | 114,000 (58,600 to 191,000) |
| 2nd most deprived quintile | 10,800 (4,560 to 19,600) | 20,500 (10,000 to 35,100) | 61,000 (30,700 to 103,000) | 31,300 (14,600 to 54,400) | 92,400 (45,800 to 157,000) |
| 3rd most deprived quintile | 10,400 (4,370 to 18,900) | 20,000 (9,640 to 34,500) | 61,700 (30,700 to 105,000) | 30,300 (14,200 to 52,900) | 92,200 (45,000 to 158,000) |
| 4th most deprived quintile | 9,000 (3,750 to 16,500) | 17,500 (8,230 to 30,400) | 55,600 (27,000 to 95,900) | 26,500 (12,200 to 46,700) | 82,200 (39,400 to 142,000) |
| Least deprived quintile | 8,860 (3,320 to 17,100) | 16,700 (7,400 to 30,600) | 53,600 (24,900 to 96,000) | 25,600 (10,800 to 47,600) | 79,200 (36,100 to 143,000) |
| By sex | | | | | |
| Female | 27,500 (11,100 to 50,800) | 55,300 (26,000 to 96,500) | 176,000 (85,300 to 302,000) | 82,700 (37,700 to 147,000) | 260,000 (124,000 to 448,000) |
| Male | 24,900 (11,000 to 44,600) | 44,900 (22,400 to 75,600) | 132,000 (66,800 to 222,000) | 69,800 (33,500 to 120,000) | 202,000 (101,000 to 340,000) |
| By disease | | | | | |
| Asthma incidence | 20,100 (9,640 to 41,700) | 47,300 (22,700 to 99,300) | 137,000 (66,000 to 286,000) | 67,400 (32,400 to 141,000) | 204,000 (98,400 to 429,000) |
| COPD severity | 26,600 (2,020 to 64,700) | 37,200 (2,780 to 93,900) | 104,000 (7,720 to 266,000) | 63,800 (4,800 to 158,000) | 168,000 (12,500 to 424,000) |
| LRTI event rate | 3,970 (843 to 8,860) | 11,400 (2,410 to 25,700) | 53,100 (11,100 to 122,000) | 15,400 (3,260 to 34,700) | 68,600 (14,400 to 156,000) |
| Health expenditure change ($millions in 2021 AUD) compared with business as usual | | | | | |
| Total | -1,680 (-3,040 to -714) | -2,130 (-3,700 to -995) | -4,690 (-8,250 to -2,230) | -3,810 (-6,690 to -1,730) | -8,490 (-14,800 to -3,910) |

| | | | | | |
|---|---|---|---|---|---|
| Income gain ($millions in 2021 AUD) compared with business-as-usual | | | | | |
| Income gain | 2,080 (788 to 3,980) | 3,780 (1,730 to 7,230) | 10,300 (5,100 to 20,100) | 5,890 (2,530 to 11,100) | 16,200 (7,760 to 31,400) |
